# The combination of autism and exceptional cognitive ability increases risk for suicidal ideation

**DOI:** 10.1101/2022.02.17.22271086

**Authors:** Lucas G Casten, Taylor R Thomas, Alissa F Doobay, Megan Foley-Nicpon, Sydney Kramer, Thomas Nickl-Jockschat, Ted Abel, Susan Assouline, Jacob J Michaelson

## Abstract

Autism with co-occurring exceptional cognitive ability is often accompanied by severe internalizing symptoms and feelings of inadequacy. Whether cognitive ability also translates into greater risk for suicidal ideation is unclear. To investigate this urgent question, we examined two samples of high-ability individuals with autism for factors that were predictive of suicidal ideation. In the first sample (N=1,074 individuals seen at a clinic specializing in gifted/talented youth), we observed a striking excess of parent-reported suicidal ideation in autistic individuals with IQ*≥*120 (OR=5.9, *p* = 0.0007). In separate sample of N=1,983 SPARK participants, we confirmed higher rates of suicidal thoughts compared to non-autistic children from the ABCD cohort (OR=6.8, *p <* 2.2 *×* 10^*−*16^), and further that autistic children with suicidal thoughts had significantly higher cognitive ability (*p <* 2.2 *×* 10^*−*16^) than those without. Elevated polygenic scores (PGS) for cognitive performance were associated with increased suicidal thoughts (*Z* = 2.16, *p* = 0.03), with PGS for educational attainment trending in the same direction (*Z* = 1.4, *p* = 0.17). Notably, similar results were found in parents of these autistic youth, where higher PGS for educational attainment was associated with increasing thoughts of suicide (Z=2.28, p=0.02). Taken together, these results suggest that on a phenotypic and genetic level, increasing cognitive ability is an unexpected risk factor for suicidal ideation in individuals diagnosed with, or at risk for autism.

## 1 Introduction

Autism is a highly genetic neurodevelopmental condition (heritability of *>* 80% [1]) affecting an estimated 1 in 44 children in the U.S. every year [2]. Research into the biology underlying autism has primarily focused on the core symptoms defined by the DSM [3]: social communication challenges and restricted or repetitive behaviors. However, less is known about the genetic factors influencing other mental health comorbidities in autism, the most alarming being suicidal ideation and death by suicide.

Previous studies of autism found profoundly increased rates of suicide and depression [4], [5], [6], [7]. The rate of death by suicide is 7.5 times higher in autistic people than those without an autism diagnosis [4]. The increased suicide rate in autism may be partially attributable to a broad increase in depressive symptoms, as autistic people have been shown to have a 4-fold increase in lifetime rates of depression [7]. Furthermore, this mental illness burden may be exacerbated by exceptional cognitive ability: in other work, we found that children with a exceptional IQ (*≥* 120) and autism have greater feelings of inadequacy and internalizing problems compared to autistic individuals with average IQ [8]. These findings contrast with findings in non-autistic cohorts, where large population studies have found high IQ is a protective factor against suicide death [9, 10], suggesting that the relationship between intelligence and suicide-related traits may vary across diagnostic boundaries.

There is evidence that suicide risk is partly genetic in nature, with heritability estimates ranging from 17-55% [11]. Some evidence for potential mechanistic overlap between the biology of suicide and of autism comes from a study that identified mutations in a well-known autism gene, *NRXN1*, as increasing risk for suicide [12]. Furthermore, increased polygenic risk for autism, in undiagnosed individuals, was found to be positively associated with suicidal thoughts, lending further evidence to potential shared biological mechanisms between suicide and autism [13]. Although connected to autism through these modes of genetic risk, these studies were completed in non-autistic samples, and it is unknown how these factors might also contribute to the burden of suicidal thoughts and suicide seen in autism.

This previous work points to several key questions, which we aim to address in this study: do children with autism show increased signs of suicidal ideation compared to peers? Are suicidal thoughts related to elevated cognitive ability in children with autism? What modes of genetic risk are associated with depressive and suicidal traits in children with autism? In seeking to answer these questions, we assembled evidence from multiple samples: a clinical sample enriched for high academic achievers with neurodevelopmental conditions [8], the Adolescent Brain Cognitive Development study (ABCD, [14]), a general population sample, and SPARK - a nationwide study of autism [15]. Answering these questions will offer new insight into context-dependent risk factors and the interplay and trade-offs between intelligence, neurodiversity, and mental illness, while also drawing attention to clinically meaningful subgroups most at risk for suicide. To our knowledge, this is the first study to examine the relationship between autism, exceptional cognitive ability, suicidal thoughts, and genetics in multiple large samples of children.

### 1.1 Language choices

Many autistic self-advocates prefer identity-first language (i.e., autistic individuals), and some autistic individuals and their families prefer person-first language (i.e., individuals with autism). We recognize the validity of the arguments behind both of these preferences, and have chosen to use identity-first language for this paper.

## 2 Methods

### 2.1 Clinical sample

Over a ten-year period (2009-2019), data from 1,254 evaluations from a university-based clinic that specializes in the assessment and counseling of gifted and twice-exceptional students were recorded [8]. Some individuals were evaluated more than once, only data from the first evaluation was analyzed leading to a final N=1074. Full Scale IQs from the Wechsler family of IQ tests [16] in this sample ranged as high as 158, and small handful of the clients evaluated at the clinic had very low IQs (e.g., 55). The mean IQ for the sample was 116.9 (SD of 14.5), median IQ was 117. All participants provided informed consent for their data to be used in research studies, and this study was approved by the University of Iowa IRB (IRB 202002251). Sample demographics are shown in Table 1.

**Table 1.** Cohort demographics.

| Sample | Max sample size | Mean age (SD) | Male (%) | White (%) | Multiracial (%) | Hispanic (%) | Black (%) | Asian or Pacific Islander (%) | Native American (%) | Other race reported (%) | No race reported (%) |
| --- | --- | --- | --- | --- | --- | --- | --- | --- | --- | --- | --- |
| clinical | 1054 | 10.2 (3.7) | 68.4 | 79.6 | 3.3 | 2.1 | 0.4 | 3.3 | 0.1 | NA | 11.2 |
| SPARK | 6766 | 11.4 (3.2) | 78.2 | 70.0 | 8.0 | 15.0 | 3.9 | 1.5 | 0.1 | 0.8 | 0.7 |
| ABCD | 11878 | 10.9 (0.7) | 52.5 | 52.0 | 9.4 | 20.2 | 15.0 | 2.0 | 0.3 | 0.6 | 0.5 |

### 2.2 SPARK cohort

SPARK [15] is an autism study of nearly 300,000 individuals in the United States. Parents of children with autism participating in SPARK were invited to complete the Child Behavior Checklist (CBCL [17]) online through the base SPARK study and a research match, data was pooled together from SPARK phenotype V7 release and another recontact study involving SPARK participants. There was complete CBCL data available for 6,766 children in SPARK, 4,171 of which were between the ages of 8 to 15 (subset used for comparison to ABCD cohort), and 1,982 had quality control passing genotype and behaviorally-predicted IQ data. The Research Match study was approved by the University of Iowa IRB (IRB 201812788) and SPARK is approved by the Western IRB (IRB 20151664). All participants provided informed consent. Sample demographics are shown in Table 1.

### 2.3 ABCD cohort

The ABCD cohort [14] is a typically-developing cohort, which was recruited regardless of neuropsychiatric conditions, meant to represent a more general population sample than case/control cohorts. Release 3 data were used, CBCL data was available on 11,878 children. Z-scaled NIH toolbox full composite scores [14] were the proxy used for IQ in this cohort, and these scores were available for 4,999 children. Sample demographics are shown in Table 1.

### 2.4 Measures of suicidality

Data for suicidal ideation from the clinical sample was taken from the clinical intake forms completed by the parents of the children, then merged with data about diagnosis and clinical IQ test scores completed by the licensed psychologist who assessed the child. Data for depressive symptoms and suicidal ideation for the SPARK and ABCD cohorts came from the CBCL [17]. The CBCL [17], a parent-report tool for assessing emotional and behavioral problems in children ages 6-18, is a well-validated questionnaire consisting of nearly 200 items. CBCL items can be combined to yield Diagnostic and Statistical Manual of Mental Disorders (DSM) subscales of clinical relevance. The DSM depressive problem scale was used as a quantitative measure of depression and clinical threshold cutoff was set as scoring *≥* the 95 percentile [17]. In order to make cohorts more comparable, only children aged between 8 and 15 years old from SPARK with complete data were included in the statistical analysis reported in Results section 3.1 and Fig. 1, reducing the effective sample size to 16,017 children. Of those, 4,171 of those children were SPARK participants, all of whom had Autism Spectrum Disorder (ASD), and 200 children with ASD from ABCD. Analyses seen in Fig. 3 used a total of 6,381 children (1,409 from SPARK who all had ASD, and 82 with ASD from ABCD). All 1,982 SPARK children with complete CBCL, IQ, and genetic data were included in later analyses not using the ABCD cohort [14].

**Fig. 1.**
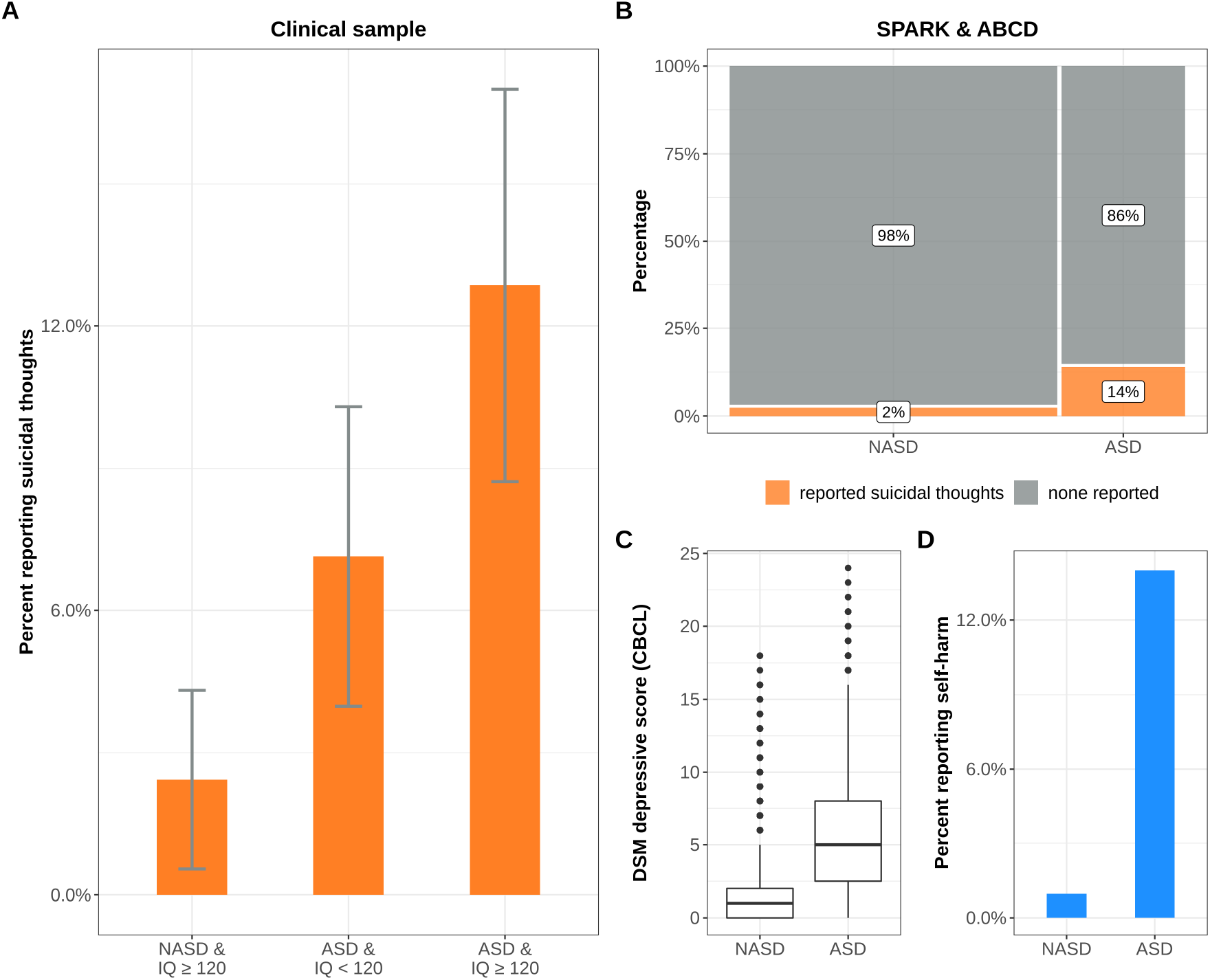
Suicidalthoughts are increased in autistic children. ASD = autistic children. NASD = non-autistic children. A. Rate of parent-reported suicidal thoughts in children seen at a clinic specializing in clinical assessment of children with exceptional cognitive ability ± 95% CI from 10,000 bootstrap samples. B. Rate of suicidal thoughts (CBCL item 91) in children in larger population samples. C. Comparison of scores on the CBCL’s DSM depressive scale. D. Reported self-harm (CBCL item 18) in children with and without ASD.

**Fig. 2.**
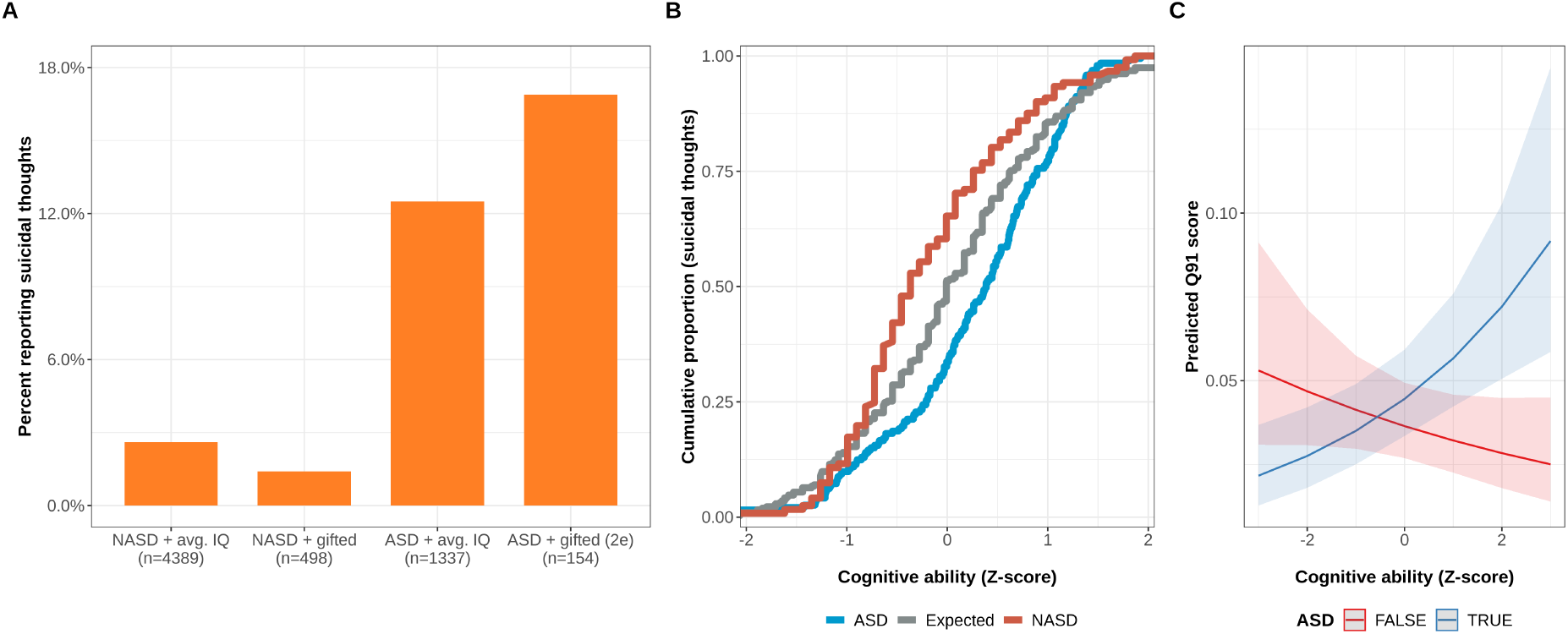
ASD and IQ interact to increase suicidal thoughts. A. Suicidal ideation in children from SPARK and ABCD, grouped by ASD status and whether their IQ proxy scores are in the top decile. B. Cumulative proportion of suicidal thoughts and their relation to Z-scaled cognitive ability scores for: ASD, NASD, and the expected rate based permutations of the complete data set. C. Predictions of suicidal thoughts from a generalized linear model trained on the SPARK and ABCD data.

**Fig. 3.**
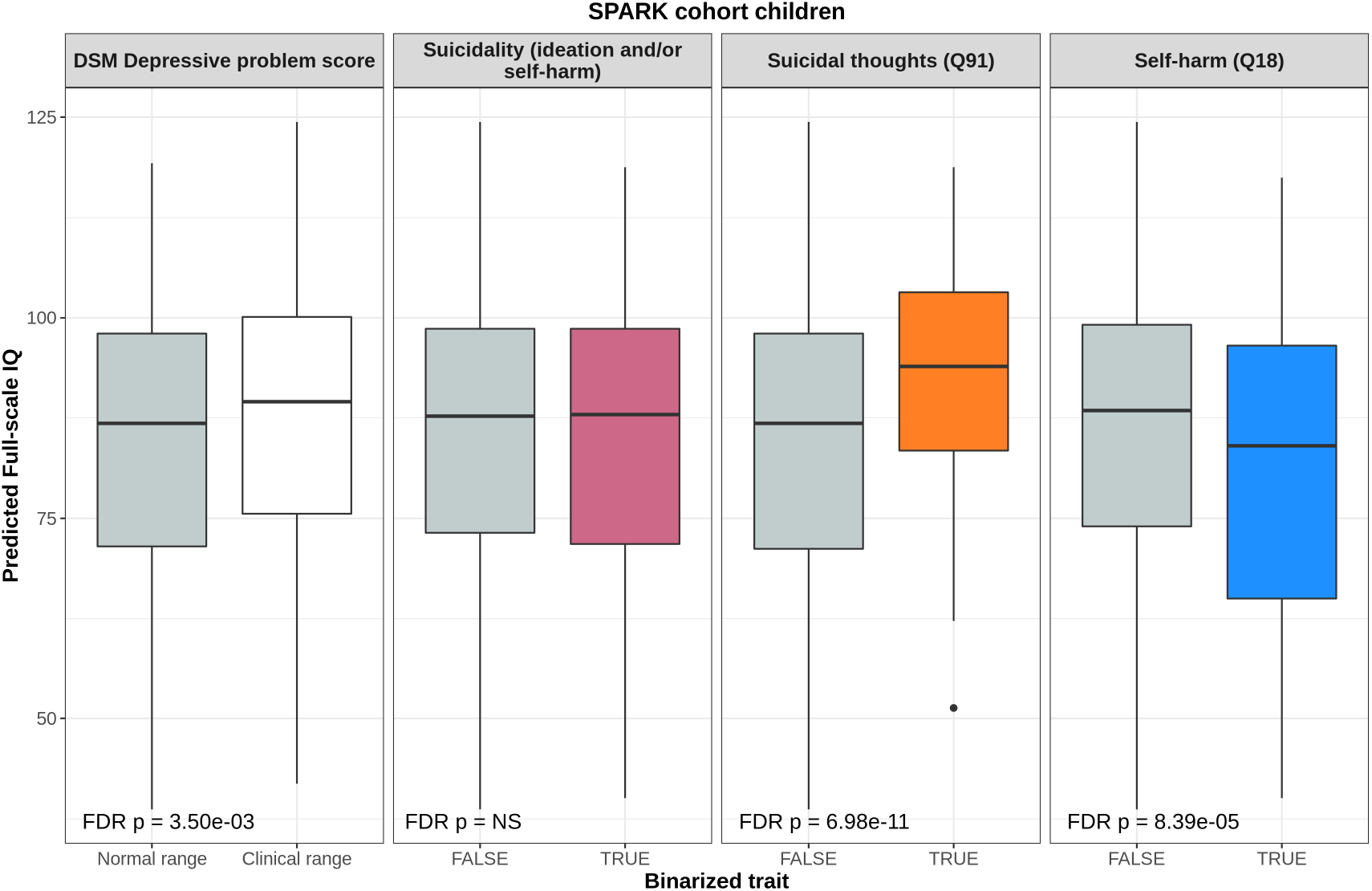
Imputed full-scale IQ scores are related to depressive phenotypes. Distributions of behaviorally-predicted IQ score split by depressive phenotypes in autistic children from SPARK. FDR-corrected p-values from Wilcox tests are reported in the bottom of each boxplot.

### 2.5 Behaviorally predicted IQ in SPARK

An imputed IQ score [18] was used because very few individuals had clinically-derived IQ scores, CBCL, and genotype data. A total of 1,982 SPARK participants had complete imputed IQ scores, CBCL, and genotype data. These predicted full-scale IQ scores [18], obtained from for SPARK phenotype v7 release, were used for the analyses depicted in Fig. 3. Briefly, these scores were predicted using a number of behavioral and autism diagnosis data (e.g. language level and repetitive behaviors), and internal validation showed these scores to be strongly correlated with clinically measured IQ (N = 2.162, Pearson’s *r* = 0.76, *p <* 2.2 *×* 10^*−*16^).

### 2.6 Genotype quality control and imputation

Genotype data from SPARK Version 3 Freeze (2019) and Version 4 (2020) were merged using PLINK ([19]). Merged genotypes were then lifted from hg38 to hg19 using the LiftOver tool [20]. The merged genotypes included 43,209 individuals and 616,321 variants and were then quality controlled using the “BIGwas” quality control pipeline [21]. The default parameters were used, except for skipping Hardy-Weinberg tests and including the flag due to the SPARK cohort being family-based and not a general population sample. The pre-QC annotation step removed 21 variants (N = 616,299 variants remaining). The SNP QC step removed 101,600 variants due to missingness at a threshold of 0.02 (N = 514,699 variants remaining). The sample QC step removed 1,114 individuals due to missingness, 67 individuals due to heterozygosity, and 176 due to duplicates (monozygotic twins). An additional 9,533 individuals were removed due to genetic ancestry from principal component projections (N = 32,422 individuals remaining). The QC’d set of N = 514,699 variants and N = 32,422 individuals were then imputed to the TopMed [22] reference panel using the Michigan Imputation Server [23] with the phasing and quality control steps included and to output variants with imputation quality *r*^2^ ¿ 0.3. After the genotype imputation, the variants were filtered to only the HapMap SNPs (N = 1,054,330 variants) with imputation quality *r*^2^ *≥* 0.8 using bcftools [24]. Next, they were lifted over from hg38 to hg19 using the VCF-liftover tool (https://github.com/hmgu-itg/VCF-liftover) and the alleles normalized to the hg19 reference genome. Finally, the files were converted to PLINK binary files [19] with N = 1,018,200 final variants, these files were used for the PGS calculations.

### 2.7 PGS calculations

Polygenic scores were calculated using LDpred2 [25] and the “bigsnpr” package [26]) in R [27]). Because SPARK is family-based, an external LD reference based on 362,320 European individuals of the UK Biobank (provided by the developers of LDpred2) was used to calculate the genetic correlation matrix, estimate heritability, and calculate the infinitesimal beta weights. Polygenic scores were calculated from the following genome-wide association studies performed by the Psychiatric Genomics Consortium: ADHD (2019) [28], autism (2019) [29], and major depression (2019) [30], bipolar disorder I (2021) [31], and schizophrenia (2021) [32]. Polygenic scores for the cognitive ability traits: cognitive performance and educational attainment were calculated using GWAS summary statistics provided by the Social Science Genetic Association Consortium [33].

### 2.8 PGS regression

Polygenic score main effects were obtained by linear modeling using PGS and covariates to predict 4 different depressive and suicide-related phenotypes from the CBCL. Covariates included were: age in months, designated sex at birth, and total DSM problems from the CBCL minus the DSM depressive problems. Generalized linear models with the ‘quasipoisson’ error distribution were used to more appropriately model the skewed count phenotype data. The *glm* function in R [27] was used for all linear modeling. An example regression equation is shown below, where *Y* _i_is the phenotype being tested, e.g., score count of DSM depressive problems from the CBCL.

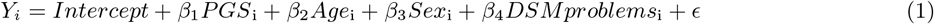

## 3. Results

### 3.1 Suicidal thoughts are associated with autism and higher intelligence

Twice-exceptional children, i.e., those with autism and at least one IQ subscale score *≥* the 90th percentile, showed the highest rate of suicidal thoughts, at 12.9%, compared to only 2.4% in IQ matched controls - demonstrated in Figure 1A (odds ratio = 5.9, odds ratio 95% CI=1.9-18.3, *p* = 0.0007). These results were consistent with those obtained from the Child Behavior Checklist (CBCL) in independent large cohorts, where nearly 14% of the 4,371 children with an autism diagnosis were reported to express thoughts of suicide, compared to 2% out of the 11,678 non-autistic children from ABCD (Fig. 1B, odds ratio=6.8, odds ratio 95% CI=5.8-7.9, *p <* 2.2 *×* 10^*−*16^). Related problems, like depressive symptoms (Fig. 1C, median sample difference = 4, 95% CI = 3.99-4, *p <* 2.2 *×* 10^*−*16^) and self-harm were also significantly increased in autism (Fig. 1D, odds ratio=16.9, odds ratio 95% CI=13.7-20.9, *p <* 2.2 *×* 10^*−*16^). There was a significant positive interaction between ASD and IQ, suggesting the relationship between IQ and suicidal thoughts is dependent on whether a child has autism or not (Fig. 2, *Z* = 3.47, FDR *p* = 0.002).

Taken together, these results from multiple large samples point toward elevated rates of suicidal thoughts in autism overall, with the highest rates being found in “twice-exceptional” or 2e individuals, i.e., those autism and exceptional cognitive ability.

### 3.2 Cognitive ability is associated with depressive phenotypes in autism

To determine the extent to which cognitive ability is associated with depressive symptoms in autism, autistic children were grouped by binarized depressive and suicidal phenotypes (e.g., suicidal thoughts vs. none reported), then tested for group differences with Wilcox tests in an imputed IQ score that has been previously described [18]. Results are illustrated in Fig 2.

Predicted IQ [18] was most significantly related to suicidal thoughts in autism: children with reported suicidal thoughts (CBCL item 91), had a mean predicted IQ score 8 points above those without reported suicidal thoughts (median sample difference = 7.2, 95% CI = 5.1-9.4, FDR *p* = 7 *×* 10^*−*11^). Conversely, autistic children and reported self-harm (CBCL item 18, which may be sensitive to a wide range of self-harm behaviors, some not associated with suicidal ideation) had significantly lower predicted IQ scores, by an average of *>* 5 points (median sample difference = -4.9, 95% CI = -2.6 - -7.2, FDR *p* = 8 *×* 10^*−*5^). This result is perhaps intuitive for clinicians and researchers who work in neurodevelopmental disabilites, and are familiar with self-harm behaviors such as banging the head against the wall or furniture, that are associated with intellectual disabilities (ID), repetitive behaviors, and other behaviors that are unlikely to be related to suicidal intent. The way CBCL item 18 is worded, “deliberately harms self or attempts suicide”, likely makes it sensitive to endorsement based on this kind of compulsive self-harm, in addition to self-harm that is more likely to be related to suicidal ideation. Notably, the binarized suicidal ideation trait (i.e., the sum of CBCL items 18 and 91) did not show any significant differences in predicted IQ, presumably due to the opposing effects of each constituent item cancelling the other out. Together, these observations warrant caution when combining items 18 and 91 of the CBCL into a “suicide score” because they may give misleading results, especially when applied within a neurodevelopmental sample. These findings are also our justification for considering items 18 and 91 separately throughout our analyses. Children who scored at or above the 95th percentile for DSM depressive problems scale (meeting the clinical problem cutoff) on the Child Behavior Checklist (CBCL) had significantly higher predicted full-scale IQ by an average of 2.5 points (median sample difference = 2.4, 95% CI = 0.8-4, FDR *p* = 0.0035). These results are in agreement with the other results relating to suicidal thoughts (Figure 1A), and together demonstrate that increasing cognitive ability is linked to a broader depressive phenotype in the context of autism. In sum, across multiple samples we find higher cognitive ability in autism to be associated with increased suicidal thoughts (CBCL item 91), increased rates of clinically-relevant depression (CBCL DSM depressive scale), and less self-harm behavior (CBCL item 18).

### 3.3 Polygenic estimators of cognitive ability predict suicidal thoughts and depressive symptoms

The final analysis aimed to determine if the observed relationship between cognitive ability and suicidal thoughts had support at the biological level. Polygenic scores (PGS) for a variety of cognitive and psychiatric traits were computed with LDpred2 [25], then tested for association with phenotypes using generalized linear models (GLM) with a quasipoisson error distribution. Effects of overall DSM problems (sum of all DSM scale scores except the DSM depression score), sex, and age were accounted for by inclusion as covariates in the GLM. Model coefficients for the PGS ± 95% confidence intervals are depicted in Figure 4.

**Fig. 4.**
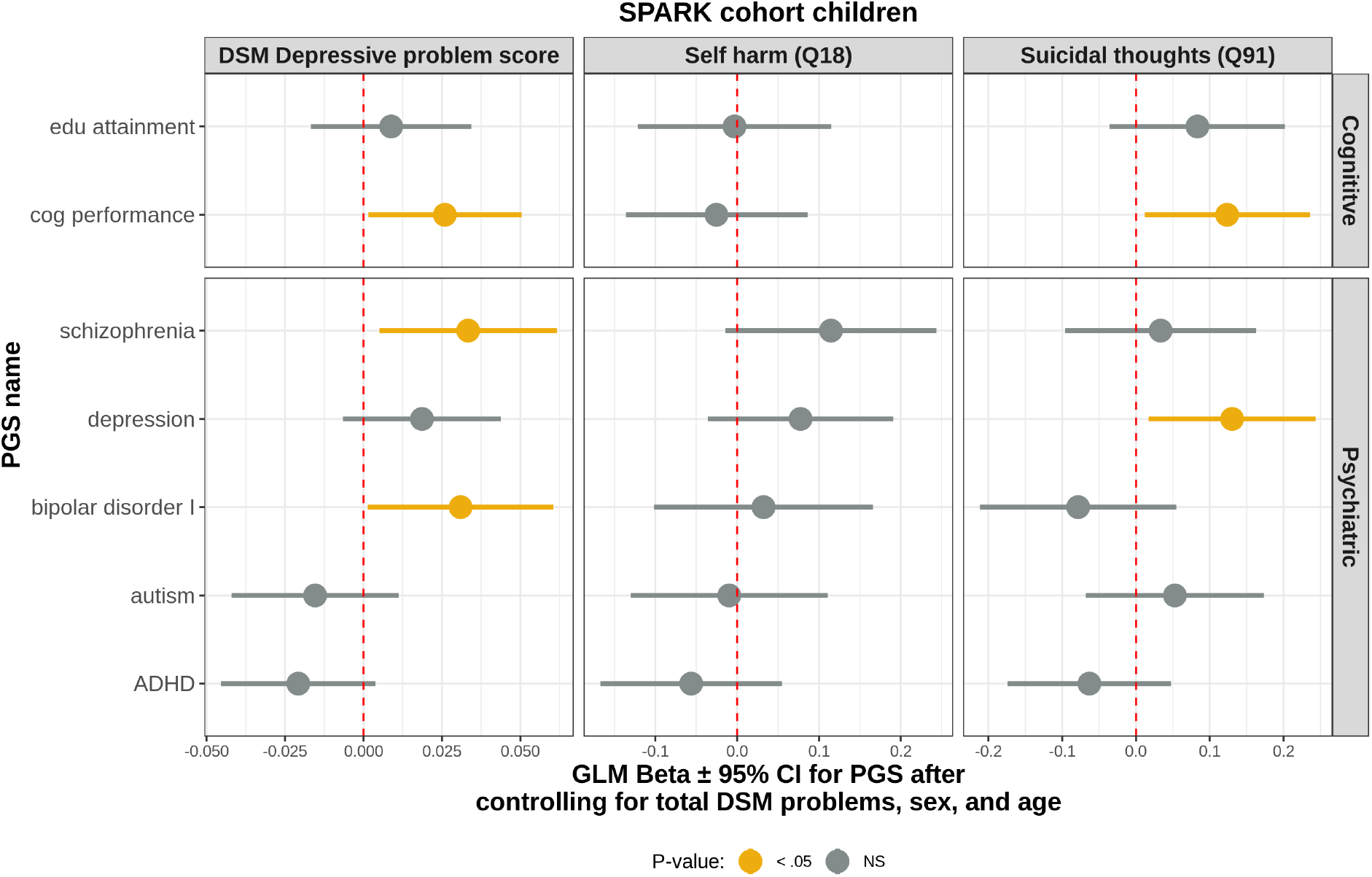
PGS for cognitive ability and depression are associated with increased suicidal thoughts. Results from generalized linear models predicting the depressive phenotype of interest. The coefficient for the PGS model term ± 95% confidence interval are indicated. Total DSM problems (except DSM depressive problems), binary sex, and age in months were included as covariates in all models.

PGS for cognitive performance was positively associated with DSM depressive problems. Higher PGS for cognitive performance [33] was associated with increased depressive symptoms in children with ASD (*Z* = 2.26, *p* = 0.02). Similarly, elevated cognitive performance PGS was positively associated with suicidal thoughts (CBCL item 91; *Z* = 2.16, *p* = 0.03). PGS for major depression was also signficantly associated with suicidal thoughts (*Z* = 2.25, *p* = 0.02). Notably, similar results were found in the parents of autistic children when examining items from the Adult Self Report (ASR) (Supplemental Fig. 1). PGS for educational attainment [33] was significantly associated with suicidal thoughts in parents of autistic children (ASR item 91; *Z* = 2.28, *p* = 0.02). Together, these results suggest that genetic propensity for higher cognitive ability increases risk for suicidal thoughts in both autistic probands and their parents.

## 4 Discussion

The main contribution of this study is in drawing attention to the significantly increased risk of suicidal ideation faced by members of a “double minority” defined by exceptional cognitive ability and autism. Our analyses demonstrate that increasing IQ switches from a protective factor in non-autistic individuals to a risk factor for suicidal ideation in autistic youth (Fig. 2). With multiple large cohorts and through a series of complementary analyses, we repeatedly demonstrated a robust link between higher cognitive ability and signs of suicidal ideation, specifically in an autistic context. There is evidence that this is at least partly genetic in nature: polygenic scores for cognitive performance and educational attainment were positively associated with thoughts of suicide in autistic youth and their parents, respectively.

Our analyses included multiple estimates of cognitive ability: the clinical sample included IQ as measured by the WISC [16], a gold standard measure, while the SPARK sample used an estimate of IQ based on a variety of behavioral items and that was calibrated against clinically-obtained IQ measures. Our genetically-informed sample in SPARK included polygenic estimators of cognitive performance and educational attainment. Regardless of how cognitive ability was estimated, we found that autistic individuals with suicidal thoughts generally have higher estimated cognitive ability than autistic peers without reported suicidal thoughts. We also found that increased cognitive ability was broadly related to increased depressive problems in autism.

Our finding that intelligence is a risk factor for suicidal ideation contrasts with previous work that has shown men with low cognitive ability are more at risk for suicide death than men with high ability [9, 34]. However, our findings are congruent with previous studies of mortality in autism that showed that higher-ability autistic adults are much more likely to die of suicide than lower-ability autistic adults [4]. Further, the rate of suicidal ideation in the SPARK sample (14%), is remarkably consistent with a previous, smaller study of suicidal ideation in autism, which found 14.5% of autistic children had suicidal thoughts [35].

To our knowledge, this is the largest genetically-informed study of risk factors for suicidal ideation and related depressive symptoms in autistic youth. It is also the first to show a biological (i.e., genetic) relationship between propensity for high cognitive ability and suicidal thoughts. We were able to directly compare suicidal ideation rates in clinical samples of autism with exceptional ability to IQ-matched non-autistic patients, showing that the presence autism is a critical factor in the relationship between cognitive ability and suicidal thoughts. This was brought into focus by the significance (*p <* 0.01) of the autism:IQ interaction term in the GLM explaining suicidal thoughts from the CBCL (see Figure 2). In non-autistic youth from the combined SPARK+ABCD sample, exceptional cognitive ability (estimated IQ at or above the 90th percentile) was shown to be a protective factor against thoughts of suicide reported via the CBCL. Strikingly, the trend was the opposite in autistic youth: those with exceptional ability were at increased risk for suicidal ideation (Fig. 2). Together, these results suggest that the effects of autism and increased cognitive are not additive when predicting suicidal thoughts as an outcome. Instead, these to factors interact.

Another unexpected finding that emerged from our analyses related to the summing of items 18 and 91 from the CBCL into a “suicide score”. We found that this summed score did not demonstrate the expected relationships with other variables of interest. Our observation that endorsement of item 18 (self harm behavior) was associated with lower indices of cognitive ability raises the possibility that this item is sensitive to compulsive and repetitive self-harm behaviors sometimes seen in individuals with an intellectual disability, and that are usually not indicative of suicidal ideation. Conversely, endorsement of item 91 (talk of suicide) is associated with higher estimated IQ. The sum of items 18 and 91 has the apparent effect of canceling out the overall association with estimated IQ. This observation would suggest that care is warranted when interpreting the meaning of item 18 with respect to suicidal intent, especially in the context of neurodevelopmental samples like SPARK.

## 5 Limitations

Despite strongly convergent evidence that increased cognitive ability is positively associated with reported suicidal thoughts in autistic children, there are a number of limitations that need to be acknowledged. All data on depression and suicidal ideation in children came from parent reports, and there are likely children in our sample with suicidal thoughts who were reported as not having any. It is likely that in the context of internalizing and depressive symptoms, including thoughts of suicide, the first person perspective provides a better signal to noise ratio, and future work should address suicidal ideation from that perspective. The current sample has several sex biases in the data: the autistic children in our samples were predominately male, while the data for parents was predominately from mothers. Previous work has shown higher-ability autistic women are at the greatest risk for suicide death [4], so a future sample where autistic females are better represented would yield key insight. Finally, we also do not know whether these early suicidal thoughts translate into greater risk for suicide attempts later in life. In the current samples, we have no data on actual suicide attempts or suicide completion. With the current single time point sample, we are unable to develop insight into the temporal and developmental effects that govern suicide risk from childhood through adolescence and adulthood.

## 6 Conclusions

Across multiple estimates of intellectual capacity (clinical, behaviorally predicted, and polygenic propensity), we found that higher cognitive ability is significantly related to more suicidal thoughts in children with autism. Specifically, autism diagnosis and cognitive ability were shown to have supralinear (i.e., interacting) effects on suicidal ideation, making twice-exceptional youth the group at highest risk for suicidal ideation. These findings build upon previous work that found suicidal ideation is significantly higher in autistic populations and offers clarity into additional mental health risk factors within autism. Future work should focus on 1) longitudinal assessment of suicide risk in youth 2) self-report of risk factors among youth and 3) increasing representation of autistic females in study samples.

## Data Availability

Data from the SPARK and ABCD cohorts are available to researchers online.
All data produced in the present study are available upon reasonable request to the authors following acceptance of the manuscript to a peer-reviewed journal.

https://www.sfari.org/resource/sfari-base/

https://nda.nih.gov/abcd

## 7 Acknowledgements

We are grateful to all of the individuals and families in SPARK, the SPARK clinical sites, and SPARK staff. We appreciate obtaining access to genetic and phenotypic data for SPARK data on SFARI Base. We are also appreciative of the individuals and families in ABCD. We are thankful to everyone involved in the clinical sample.

This work was supported by a grant from the Simons Foundation (SFARI Explorer 594788 to JJM). This work was also supported by the University of Iowa Hawkeye Intellectual and Developmental Disabilities Research Center P50 HD103556 (TA and Lane Strathearn, multi-PIs). Additional funding for this work came from the Department of Psychiatry at the University of Iowa and in part by National Institutes of Health through a Predoctoral Training Grant (T32GM008629 to LC and TT) and Research Grant DC014489 to JJM.

## 9 Figures and Tables

**Supplementary Fig. 1.**
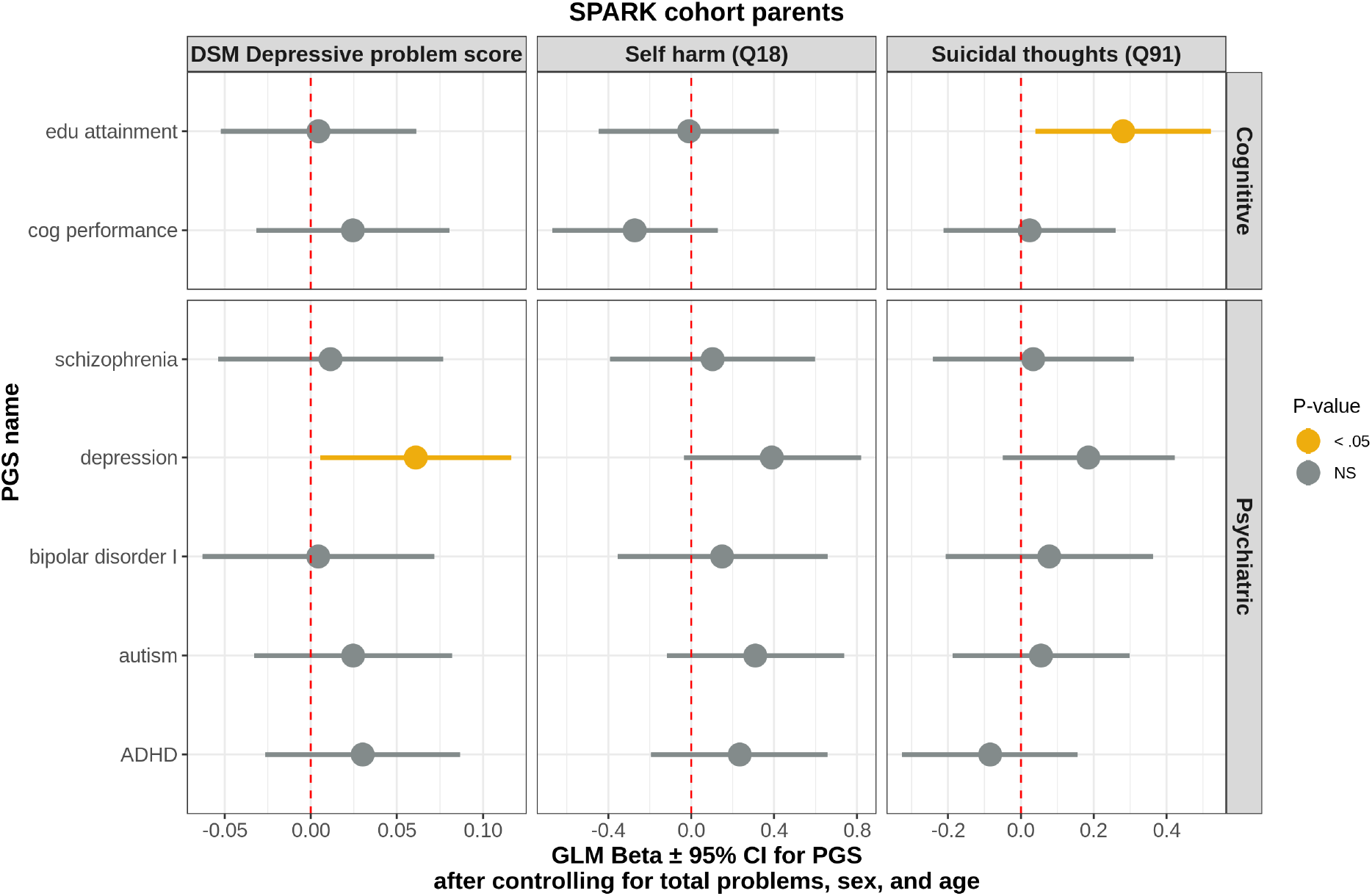
PGS for educational attainment is predictive of increased suicidal thoughts in parents. Results from generalized linear models predicting the depressive phenotype of interest. The coefficient for the PGS model term ± 95% confidence interval are plotted. Total DSM problems (except DSM depressive problems), binary sex, and age in months were included as covariates in all models.

